# Lack of children in public medical imaging data points to growing age bias in biomedical AI

**DOI:** 10.1101/2025.06.06.25328913

**Authors:** Stanley Bryan Zamora Hua, Nicholas Heller, Ping He, Alexander J. Towbin, Irene Y. Chen, Alex X. Lu, Lauren Erdman

## Abstract

Artificial intelligence (AI) is rapidly transforming healthcare, but its benefits are not reaching all patients equally. Children remain overlooked with only 17% of FDA-approved medical AI devices labeled for pediatric use. In this work, we demonstrate that this exclusion may stem from a fundamental data gap. Our systematic review of 181 public medical imaging datasets reveals that children represent just under 1% of available data, while the majority of machine learning imaging conference papers we surveyed utilized publicly available data for methods development. Much like systematic biases of other kinds in model development, past studies have demonstrated the manner in which pediatric representation in data used for models intended for the pediatric population is essential for model performance in that population. We add to these findings, showing that adult-trained chest radiograph models exhibit significant age bias when applied to pediatric populations, with higher false positive rates in younger children. This work underscores the urgent need for increased pediatric representation in publicly accessible medical datasets. We provide actionable recommendations for researchers, policymakers, and data curators to address this age equity gap and ensure AI benefits patients of all ages.

**1-2 sentence summary:** Our analysis reveals a critical healthcare age disparity: children represent less than 1% of public medical imaging datasets. This gap in representation leads to biased predictions across medical image foundation models, with the youngest patients facing the highest risk of misdiagnosis.

## Background

A growing number of studies highlight the potential for artificial intelligence (AI) to improve patient care^1,2^. A prerequisite for medical AI is the availability of high-quality datasets such as MIMIC-III^3,4^ and CheXpert^5^. The public release of these datasets attracts efforts from the machine learning community to develop new state-of-the-art algorithms, leading to advancements in medical AI, from sepsis prediction to acute critical illness management and insights for chronic diseases^6–9^.

In the United States, growing interest and development in medical AI is reflected in a year-on-year increase in the number of AI-enabled medical devices approved by the Food and Drug Administration (FDA). Yet, studies into FDA approvals reveal that fewer than 1 out of 5 devices are approved for pediatric use^10^, and ages of study participants are not reported in 81.6% of devices^11^. The neglect of age-related differences mirrors historical patterns in drug^12^ and device development^13,14^, where children suffer disparities in quality of care compared to adults^15–17^.

A growing concern in medical AI is the presence of biases in algorithmic output, driven by missing data and a legacy of inadequate care for the very populations these tools aim to help^18^. In the clinic, these algorithmic biases have the potential to harm patients, especially in high-risk applications like diagnosis and treatment planning. While algorithmic biases have been extensively studied in the context of sex and race^18^, recent studies indicate that age representation is equally important, as AI trained on adult data display age-related biases against children^19–23^. Underlying the challenging transfer of adult AI to children are fundamental differences in anatomy and physiology^24–27^, in addition to clinical differences in pediatric disease risk, presentation and management^28–35^. These stark differences suggest that pediatric data is critical in developing AI applications for children.

While the availability of public data is beneficial for developing medical AI, motivations for using public datasets vary greatly from researcher to researcher. In communities focused on a particular application, efforts tend to concentrate on a few key benchmark datasets^36,37^. In contrast, challenges attract generalist machine learning practitioners to work on novel applications. Despite the ecosystem of public datasets and challenges that help drive AI development, pediatric representation in these crucial resources and its implications remain largely unknown.

To address the gap in knowledge about pediatric representation in public data, we conduct the largest systematic review of public medical imaging datasets to date. We begin our review by analyzing data usage in an AI conference for medical imaging, where we show few studies focused on pediatric applications. We then analyze 181 public medical imaging datasets to establish this lack of research likely stems from a lack of pediatric data. Finally, we demonstrate downstream consequences of the lack of pediatric data in model development, by showing that in one case example, the use of adult-trained models results in systematically increasing false positive rates on younger children.

## Methods

### Scoping Review of Recent AI for Medical Imaging Papers

To understand patterns of pediatric representation in medical AI research, we reviewed papers accepted to the Medical Imaging with Deep Learning (MIDL)^38^ conference in 2023 and 2024. Papers are filtered for those that use structural non-invasive internal imaging data, further defined in the dataset inclusion criteria. For each paper, we recorded the dataset(s) used, and then for all datasets, we identified how the dataset was made available. Based upon this, we identified sources for datasets: datasets collections, challenges and benchmarks. More details are provided in the *Dataset Selection* section.

### Review of Public Medical Imaging Data

Given the sources of public datasets identified, we conducted a systematic review of publicly available medical imaging datasets by collecting datasets from each data source to assess pediatric representation in public data, (Figure 1a**, 1b)**.

**Figure 1.**
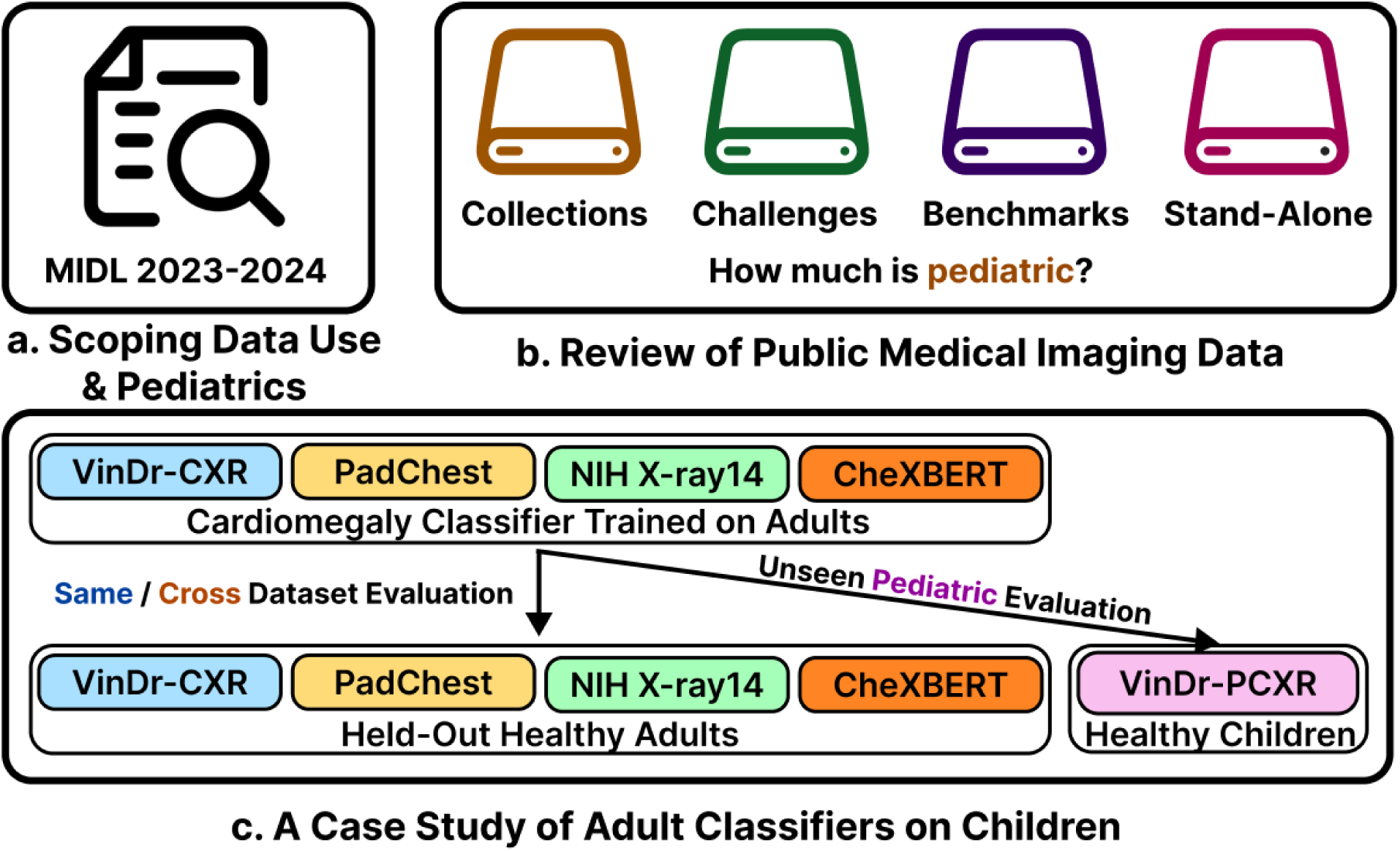
Overview of methodology. **(a)** Medical Imaging with Deep Learning (MIDL) papers are reviewed to scope usage of public datasets and presence of pediatric applications of AI for medical imaging. **(b)** Public medical imaging datasets are taken from multiple sources and reviewed for pediatric representation. **(c)** Cardiomegaly classifiers are trained on adult chest radiograph datasets and evaluated on held-out healthy adults and healthy children.

#### Dataset Inclusion Criteria

To be eligible for our systematic review, a dataset must meet the following conditions:

1. **Structural Non-Invasive Internal Imaging**. The dataset must contain data for medical imaging modalities that capture internal anatomical structures of organs and tissues and don’t require breaking the skin or physically entering the body. This strictly includes fundus imaging, ultrasound, magnetic resonance imaging (MRI), X-ray, computed tomography (CT), or their common variants (e.g., mammography, retinal OCT).
2. **Accessibility**. The dataset must be publicly downloadable and usable without requiring manual authorization from the data provider.

To avoid redundancy in downstream analyses, we excluded datasets that simply repurpose datasets already annotated. For datasets that are updated annually such as the Brain Tumor Segmentation challenge (BraTS)^39^, only the latest versions (as of January 1, 2025) containing complete and original data are included. If a dataset contains both data that falls within the inclusion criteria and data that falls outside the inclusion criteria, only the subset of data that falls within the inclusion criteria is annotated and included in subsequent analyses.

#### Dataset Annotation

For datasets meeting the inclusion criteria, we categorized how patient ages are reported and attempt to identify the proportion of pediatric patients -- patients aged 17 or younger in each dataset. We identified the following patient age reporting strategies: *patient-level*, *binned patient-level*, *summary statistics*, *task/data description* and *not documented*. In the best case, metadata files are provided containing ages for each study participant, which allows for computing the exact proportion of pediatric patients in a dataset. In *binned patient-level* reporting, patient ages are provided in age bins (e.g., 20-30 years old). If we could not infer the presence and proportion of pediatric data through available metadata files, associated publications were used to infer the presence and proportion of pediatric data through *summary statistics*. If none of the above methods were successful, we attempted to determine if there was no pediatric presence by scanning *task or data descriptions* for any clear mention or indicators of “adult-only” data in the associated publication, dataset’s official website or portal. Otherwise, a dataset was labeled as *not documenting* age.

#### Dataset Selection

To curate datasets for our systematic review, we collected and organized them by their source.

*Dataset Collections* refer to repositories curated for diverse research and practical applications, including medical imaging. For this study, we focused on clinical data repositories containing medical imaging datasets, either derived from clinical studies or explicitly released for developing machine learning algorithms. Our analysis included the following dataset collections: **(a)** The Cancer Imaging Archive (TCIA)^40^, **(b)** Stanford AI in Medicine & Imaging (AIMI)^41^, **(c)** UK BioBank^42^, **(d)** Medical Imaging and Data Resource Center (MIDRC)^43^, and **(e)** OpenNeuro^44^. For TCIA and AIMI, we focused on complete datasets strictly with patient-specific age metadata, while for UK BioBank, MIDRC and OpenNeuro, we relied on aggregated patient statistics provided by their respective portals.

*Challenges* play a pivotal role in advancing state-of-the-art machine learning, often targeting novel or domain-specific applications. Machine learning challenges for medical imaging are commonly hosted on platforms such as the Grand Challenge^45^, Kaggle^46^ and the Radiological Society of North America (RSNA)^47^ in conjunction with conferences. For this study, we annotated datasets from challenges hosted on the listed platforms, launched in the last five years (2019 to 2024).

*Benchmarks* consist of one or more pre-existing or novel datasets designed to evaluate and compare algorithm performance on specific and often well-established tasks. While numerous datasets can serve as benchmarks, we focused on three explicitly labeled as such: two benchmarks for multi-organ segmentation: Medical Segmentation Decathlon (MSD)^48^ and AMOS^49^, and one benchmark for assessing fairness in medical imaging machine learning: MedFAIR^50^.

*Well-Cited Datasets* serve as a means to identify datasets that don’t belong to a distinct dataset collection, challenge or benchmark, yet have become foundational resources in the academic community, serving as the basis for numerous significant contributions over time. To identify these datasets, we leveraged Meta’s PapersWithCode^51^, which ranks imaging datasets by citation count. We selected and annotated the top 30 datasets meeting our inclusion criteria. When aggregating datasets across dataset sources, we remove any duplicate datasets found via PapersWithCode and the other dataset sources. In analysis on well-cited datasets alone, all 30 identified datasets are used.

#### Data Breakdown by Modality and Task

To investigate if pediatric representation differs by application, we first filtered for datasets where the proportion of pediatric data can be estimated, then we categorized datasets by imaging modalities and machine learning tasks. Specifically, machine learning tasks are classified under: a) *condition classification* (e.g., pneumonia classification), b) *anatomy segmentation* (e.g., organ or tissue segmentation), c) *lesion segmentation* (e.g., tumor segmentation), d) *disease risk segmentation* (e.g., fractured rib segmentation, organs-at-risk segmentation), and e) *image enhancement* (e.g., MRI image reconstruction, synthetic image generation). To perform our analysis, we aggregated the number of medical images across datasets by imaging modality and task. As any one dataset can contain data from multiple imaging modalities, we attempted to identify what proportion of the dataset comprises each modality, and if not possible, we assumed the images are split evenly across modalities.

#### Dataset Repackaging

In our review, we observed cases in which a released dataset includes data previously released in a pre-existing public dataset. We term this phenomenon “dataset repackaging”. We identified all datasets that perform dataset repackaging. On those datasets, we annotated the original datasets they reference and extracted age-related metadata from the original dataset. As many foundation models are often developed on public datasets, we supplemented our analysis by considering patient ages in the datasets used by MedSAM, a medical foundation model.

### A Case Study of Adult Cardiomegaly Classifiers on Children

To examine how medical AI developed on adult data underperforms on children, we trained models on chest radiographs to classify cardiomegaly, a condition characterized by an abnormally large heart. (Figure 1c)

#### Data

For our experiments, we utilized five large publicly available chest x-ray datasets: **(a)** VinDr-PCXR^52^, **(b)** VinDr-CXR^53^, **(c)** NIH X-ray14^54^, **(d)** PadChest^55^ and **(e)** CheXBERT^5,56^. VinDr-PCXR is a dataset of posteroanterior (PA) view chest x-rays from children aged 0 to 10 years, collected at a major pediatric hospital in Vietnam. Healthy children with age annotations in the VinDr-PCXR dataset are used for held-out pediatric evaluation. The remaining chest x-ray datasets consist primarily of adult data and are used for training, calibration and held-out evaluation on healthy adults. The VinDr-CXR data is from two hospitals in Hanoi, Vietnam, while PadChest data is from a hospital in Barcelona, Spain. Meanwhile, the NIH X-ray14 and CheXBERT are from hospitals in the United States associated with NIH and Stanford, respectively. NIH X-ray14 is the extended version of the original NIH ChestX-ray8 dataset, while CheXBERT is the CheXpert dataset with CheXBERT-assigned labels.

#### Model

A ConvNeXt-B (89M parameters) is chosen as our base neural network architecture^57^. For each of the datasets, a network is trained for at most 50 epochs using the AdamW^58^ optimizer with a learning rate of 0.0001 to optimize the binary cross-entropy loss. During each training step, 32 random x-ray images are sampled equally from both classes, and MixUp is used for data augmentation^59^. Parameters from the epoch with the best validation set loss are kept.

#### Evaluation

The probability threshold (or operating point) for each classification network is adjusted to maximize Youden’s J statistic (defined as sensitivity + specificity - 1) on their corresponding calibration set. Each adjusted model is then used to predict cardiomegaly on the healthy children in the VinDr-PCXR dataset and healthy adults in each of the adult datasets. The proportion of false positives is reported at the dataset level and stratified by age groups. To determine statistically significant differences, bias-corrected and accelerated bootstrap is used to construct 95% confidence intervals around the false positive rate^60^. To identify patterns between age in years and false positive rate, we normalized the false positive rates by each model and computed a Pearson r correlation.

## Results

### AI research studies consistently underrepresent children

Among 46 studies from MIDL 2023-2024, only one study (2.2%) directly applied its work to pediatrics. Moreover, 85% (39/46) of studies and 35% (22/62) of cited public datasets failed to report patient ages altogether – a fundamental metadata gap that obscures representational analysis. Only one of the 46 studies used a dataset containing primarily pediatric data, and while 9 additional studies used datasets containing pediatric data, less than 5% of the patients in those datasets were children. Across MIDL studies, public datasets are used in the majority (37/46; 80%) of studies. From this, we hypothesized that limited pediatric applications may stem from a lack of public pediatric datasets, motivating a larger review of the presence of pediatric patients in public medical imaging data.

### Children are systematically underrepresented in public medical imaging data

Our comprehensive review of 181 public datasets reveals a large disparity: only 3.3% (6/181) of datasets are pediatric-only, while an additional 14.4% (26/181) of datasets contain both adult and pediatric data. Among 116 datasets with sufficient patient age information, children represent under 1% of patients in public medical imaging datasets (4.6K of 489K) – a stark contrast to their 22% share of the population in the United States^61^, where most of these patients were imaged (Figure 2a). This underrepresentation persisted across all dataset sources (Figure 2b, **Table 1**). In major collections like MIDRC^62^, TCIA^40^, and Stanford AIMI^63^, children comprised merely 1-2% patients. Other data collections such as the UK BioBank^42^ exclude pediatric patients by design. Among 59 ML challenges between 2017-2024, only two explicitly targeted pediatric applications. Even established benchmarks showed minimal pediatric presence: zero evidence of pediatric data in MSD^48^, and less than 1.1% in AMOS^49^ and 0.1% in MedFAIR^50^. Among the 30 most-cited standalone datasets, children represented just 0.8% of patients.

**Figure 2.**
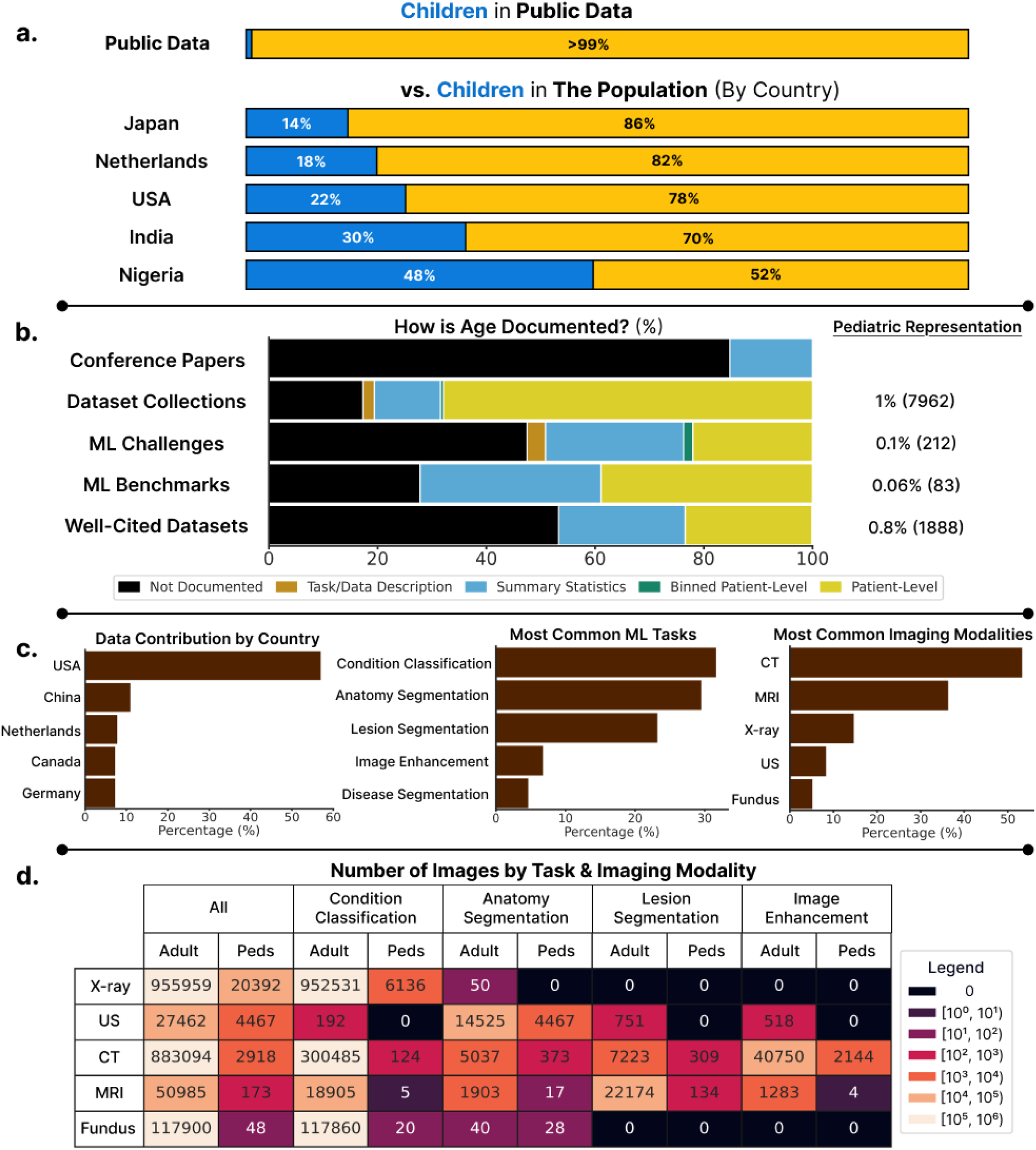
**(a) Children make up less than 1% of patients in public medical imaging datasets**. This is contrasted with the true population percentages provided by the United Nations^61^ in 5 countries selected from our dataset review. **(b) There is a lack of standardized patient age reporting in datasets released and the papers that cite them**. For each dataset source, the percentage and number of pediatric patients is reported across datasets where age is documented (right). **(c) Descriptive plots on source countries, tasks and imaging modalities in reviewed public medical imaging datasets**. Percentages provided describe the percentage of datasets that satisfies the condition. **(d) Stratifying the number of images by modality and task across 120 datasets reveals data scarcity for pediatric applications**.

**Table 1.** Summary statistics of public medical imaging datasets studied from each data source. Statistics are estimated using papers and metadata associated with each dataset. **N** denotes the number of collections, challenges, benchmarks or well-cited datasets. The remaining columns provide counts on the number of unique countries, unique institutions, unique datasets and estimated number of patients.

|  | **N** | **Countries** | **Institutions** | **Datasets** | **Patients** |
| --- | --- | --- | --- | --- | --- |
| **Dataset Collections** | 5 | 13 | 68 | 958 | 788.9K |
| **ML Challenges** | 59 | 28 | 103 | 59 | 214.3K |
| **ML Benchmarks** | 3 | 14 | 39 | 18 | 143.1K |
| **Well-Cited Datasets** | 30 | 15 | 41 | 30 | 259.3K |

The true extent to which children are underrepresented is obscured by the lack of standardized age reporting across datasets. This led to the exclusion of 63% of well-cited datasets, 83% of benchmark datasets, 51% of ML challenges and 54% of datasets in OpenNeuro, TCIA and Stanford AIMI collections.

### Stratifying by modality and task reveal many applications for which virtually no pediatric data exists

Grouping adult and pediatric images across datasets, we found little to no pediatric representation in many modality-specific tasks relative to adults (Figure 2d). When comparing by imaging modality alone, the data gap varies greatly with the smallest in ultrasound: an estimated 6 adult for every 1 pediatric ultrasound image, while larger gaps are observed for X-ray (47:1), MRI (295:1), CT (302:1) and fundus (2456:1). In our analyses, often one dataset comprises the majority of pediatric data for each modality: X-ray (70% from RSNA Pediatric Bone Age^64^), ultrasound (100% from EchoNet-Peds^20^), CT (72% from LoDoPaB-CT^65^), MRI (77% from BONBID-HIE^66^) and fundus (100% from CHASE DB 1^67^).

### Patient ages are often discarded and overlooked in data repackaging efforts

Nearly 10% (16/181) of the datasets we reviewed repackage data from other public datasets. Among datasets that repackage data, half (8/16) used public datasets that included age metadata. Yet, patient ages were discarded in the resulting datasets. Further, little attention is paid to patient ages in recent efforts to combine public datasets for foundation model development. In MedSAM, 26 public datasets are aggregated to create a dataset of nearly 1.6 million medical image-mask pairs with no information on the distribution of patient ages. Pediatric data is only present in 3 of the 26 datasets with an estimated 14 children among 8508 patients overall. For recurring competitions, pediatrics appears to be a more recent and intentional development. While BraTS^39^ exclusively targeted adults from 2012 to 2022, BraTS 2023^22^ was the first to focus on pediatric gliomas. While this indicates a shift towards recognizing the importance of pediatrics, newer pediatric datasets currently represent a small fraction over a legacy of adult-focused datasets. These observations suggest that data repackaging efforts, if not intentional about age, may lead to age-imbalanced datasets given the current data landscape.

### Pediatric data is necessary to develop pediatric-targeted AI applications as adult models are not guaranteed to transfer well to children

In the absence of tailored solutions, medical devices and drugs designed for adults may be used on children^15,16^. The untested pediatric use of medical devices and drugs introduces unknown risks to children^17,68^, and we argue similar risks exist in the new era of AI-enabled medical devices. To demonstrate hidden age biases in adult-trained AI models, we trained cardiomegaly classifiers on four adult chest X-ray datasets, and measured their false positive rate on held-out healthy adults and on healthy children from a held-out pediatric X-ray dataset (Figure 3). On healthy adults, the models did not display consistent age-related bias across adult datasets. In children, however, we found a significant age-related bias: for patients under 11 years old, false positives as patient age decreases (Pearson r=-0.928, p-value<0.0001), irrespective of the dataset the model was trained on. Additionally, we showed that correcting for differences in image contrast and quality using histogram matching fails to rescue the high false positive rates for younger children across models **(Supplementary** Figure 2).

**Figure 3.**
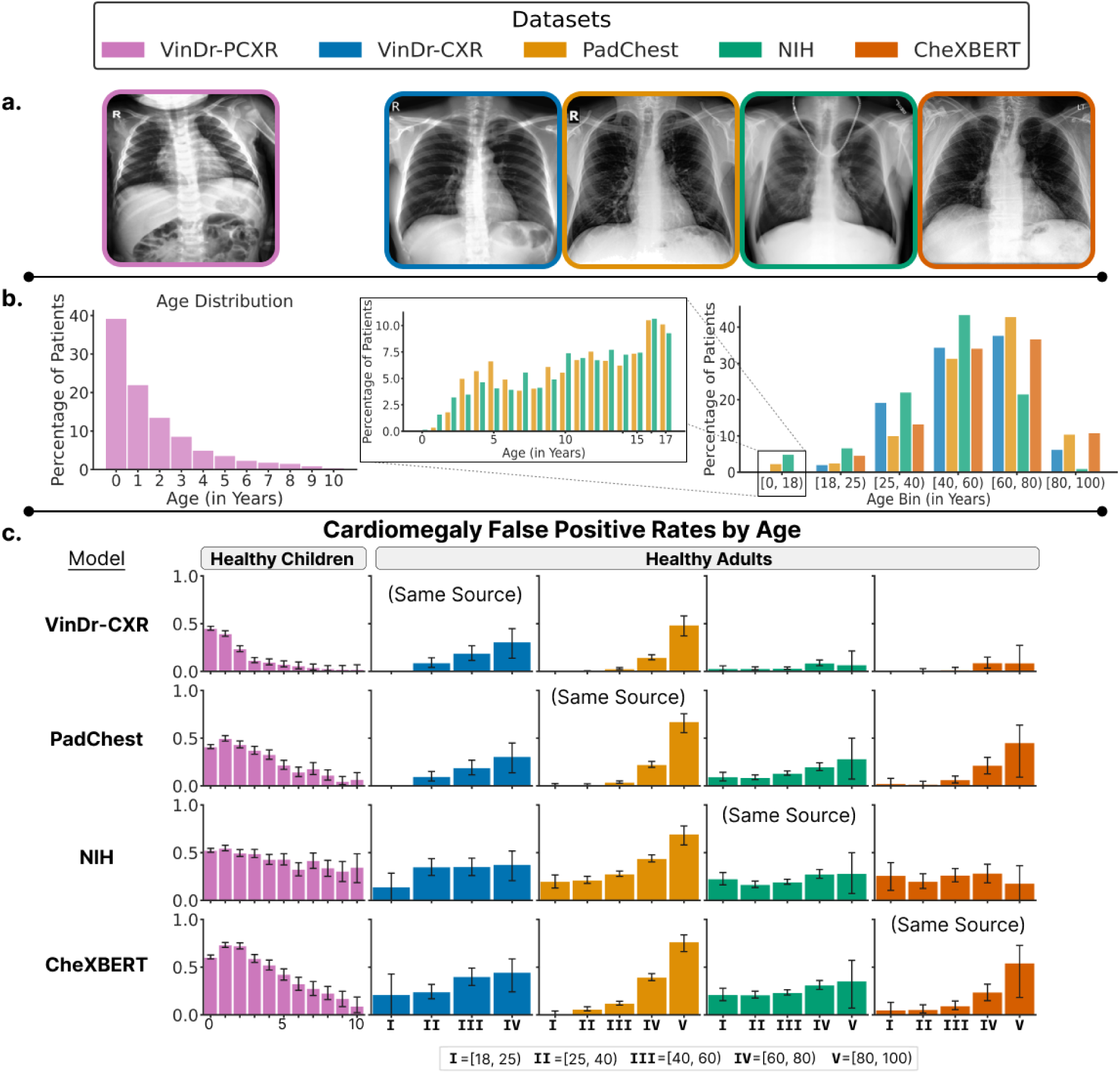
(a) Example chest x-ray image from a healthy patient in each of the chest x-ray datasets; the pediatric evaluation dataset VinDr-PCXR (**left**) and the adult-majority training datasets (**right**). **(b) Distribution of patient ages** in VinDr-PCXR dataset (**left**) and in the adult-majority datasets (**right**). For adult-majority datasets with pediatric data (i.e., Padchest, NIH), the pediatric age distribution is skewed towards older children with almost no infants at all (**middle**). **(c) Cardiomegaly classifiers trained on adult data increasingly mispredict cardiomegaly on younger children**. The y-axis represents the false positive rate, and error bars display bootstrapped 95% confidence intervals.

## Discussion

Our study reveals a critical data gap: children represent less than 1% of all patients in public medical imaging data. Only 1 of the 30 (3.3%) most cited medical imaging datasets is pediatric, implying little attention to pediatric AI historically. Only 2 in 59 (3.4%) medical imaging challenges between 2017-24 are pediatric, and only 1 of 46 (2.1%) MIDL studies in 2023-24 targets pediatrics. Altogether, these findings suggest a lack of pediatric AI applications being actively worked on in the research community, potentially driven by the pediatric data gap. In the absence of pediatric AI models, practitioners may opt for the off-label use of adult AI models. While prior work has shown that adult-trained AI algorithms can underperform on children^19–23^, we demonstrate an increasing pattern of age-related bias in chest radiographs, showing that adult-trained models increasingly mispredict cardiomegaly in young children.

To identify and prevent age-related model failure in practice, pediatric representation is crucial yet currently overlooked in publicly available medical imaging data. Beyond representation, we also identify issues with documentation: most public datasets fail to report patient ages, which limits age-related analyses and model corrections for age-related bias. Our analyses further establish that pediatric representation in public data is not only small but fragmented. Pediatric data in each modality is dominated by 1 dataset, and many tasks see virtually no pediatric representation in certain modalities.

With growing demand for larger datasets to fuel foundation model development, our findings suggest that the lack of diversity of pediatric datasets may bias dataset curators towards aggregating adult datasets, and the lack of age metadata may prevent attempts to balance data by age.

Additionally, the pediatric data gap appears to vary greatly by imaging modality with 1 pediatric image for every 6 adult images in ultrasound, compared to CT (1 for 302) and MRI (1 for 295). Since children receive more ultrasounds than CT and MRI^69–71^ compared to adults, this may suggest that differences in medical practice may contribute to the pediatric data gap. Pediatricians also employ different imaging strategies and protocols for children^23^. While scaling public pediatric data is necessary to resolve the data gap, we caution against data collection efforts that may attempt to standardize imaging practices to match that of adults, as it poorly reflects pediatric imaging in practice.

We acknowledge the systematic barriers that discourage pediatric data curation and release. Developing medical devices and drugs for pediatric use faces unique economic and regulatory hurdles compared to medical devices and drugs for adult: the rarity of some pediatric pathologies lead to small sample sizes, poor patient recruitment and high development costs^72,73^ and pediatric clinical trials requiring greater standards for consent, safety and efficacy^73–76^. While changes in public policy have greatly encouraged pediatric drug and device development, similar policy changes have yet to reach medical AI and encourage pediatric data collection^77–79^. An encouraging sign is the formation of new initiatives to support pediatric AI equity^80,81^. However, we believe that closing the gap will require collective efforts from diverse stakeholders.

### A solution to the pediatric data gap will not arise naturally

We urge the broader ML for health community to take action. For healthcare institutions and researchers, we encourage greater collaboration and initiatives to collect, prepare and release AI-ready pediatric data to the public; to support the development of pediatric AI applications. We emphasize the necessity of age representation from infancy to adulthood and the reporting of ages at the patient-level. Greater age representation and transparency will help to identify age ranges where AI models are most effective, leading to improved safety beyond pediatric cases. Building on public pediatric data releases, we encourage the creation of challenges and benchmarks to leverage the benefits of community-driven algorithm development and spread awareness on the necessity for pediatric AI. For policy-makers, we hope for policy changes that lower barriers to and encourage pediatric data collection and sharing. In addition, we argue for greater scrutiny in the pediatric evaluation of AI applications, focusing on precision in targeted age groups and sufficient representation across ages of human development.

### Limitations

We acknowledge two limitations of our study. First, there may exist pediatric datasets that were not captured in our systematic dataset search, particularly in data platforms without centralized curation such as Zenodo and Harvard Dataverse. Our analysis, however, captures key pathways in which datasets may be found by ML practitioners. Lastly, poor age reporting among public datasets led to the exclusion of many datasets in our analysis. Although pediatric data may exist among the excluded datasets, our findings suggest they may only represent a small portion.

## Conclusion

Despite making up 30% of the world’s population^61^, children are almost invisible in the datasets driving advancements in AI for medical imaging, constituting less than 1% of public medical imaging datasets. The glaring underrepresentation of pediatric patients and the poor reporting of patient ages in public medical datasets are critical oversights that limit the development of safe AI for pediatric use. This work not only quantifies the extent of this problem but demonstrates the potential risks of using adult-only AI models on children. By generating, sharing, and developing methods using pediatric medical data, we can ensure that the benefits of AI-driven healthcare innovations are equitably distributed across all age groups.

## Data & Code Availability

All data and code to reproduce analyses in this paper are available on GitHub at https://github.com/stan-hua/ped_vs_adults-cxr. All annotations for our dataset review are provided in the attached **Supplementary File 1**.

## Data Availability

Annotations for all datasets reviewed and code to reproduce cardiomegaly experiments are made available in the associated GitHub repository: https://github.com/stan-hua/ped_vs_adults-cxr. Specifically, all dataset annotations can be found in the following file: https://github.com/stan-hua/ped_vs_adults-cxr/blob/main/data/metadata/open_data_metadata.xlsx

## Acknowledgements

Data processing, data analysis was performed at the High-Performance Computing Facility, Centre for Computational Medicine, The Hospital for Sick Children, Toronto, Canada. This research was enabled in part by support provided by Compute Ontario (computeontario.ca) and the Digital Research Alliance of Canada (alliancecan.ca). ***MIDRC.*** The imaging and associated clinical data downloaded from MIDRC (The Medical Imaging and Data Resource Center) and used for research in this publication was made possible by the National Institute of Biomedical Imaging and Bioengineering (NIBIB) of the National Institutes of Health under contract 75N92020D00021 and through The Advanced Research Projects Agency for Health (ARPA-H). The content is solely the responsibility of the authors and does not necessarily represent the official views of the National Institutes of Health. ***Stanford AIMI***. This research used data provided by the Stanford Center for Artificial Intelligence in Medicine and Imaging (AIMI). AIMI curated a publicly available imaging data repository containing clinical imaging and data from Stanford Health Care, the Stanford Children’s Hospital, the University Healthcare Alliance and Packard Children’s Health Alliance clinics provisioned for research use by the Stanford Medicine Research Data Repository (STARR).

## Supplementary Methods

### A Case Study of Adult-Trained Cardiomegaly Classifiers on Pediatric Patients

#### Dataset Standardization

To standardize the adult datasets, the following steps are taken:

i. Exclude pediatric patients aged 17 and under, if any.
ii. Exclude data points without findings annotated or with particularly invalid age annotation.
iii. Keep only PA view images.
iv. Sample one image per patient, for patients with data from multiple visits, in priority of: **a)** having valid age annotated, and **b)** having cardiomegaly.
v. Exclude patients with findings other than cardiomegaly and patients with uncertain cardiomegaly findings.

Each of the four adult-only datasets are then split independently into a training set, calibration set and held-out set of healthy adults. To create the held-out set, 10% of healthy adults are sampled from each age bin in [18, 25, 40, 60, 80, 100], with the exception of VinDr-CXR where 20% was sampled due to fewer adults having valid age annotations. The calibration set is curated from 10% of the chest x-rays sampled from patients with cardiomegaly, patients without any findings, and patients with a finding other than cardiomegaly. After which, data from patients with a finding other than cardiomegaly are removed. Of the remaining data, 75% is used for training, while the other 25% is used for validation **(Supplementary Table 1)**. All images are loaded in RGB channels and resized to (224, 224). For each of the four adult datasets, we computed dataset-specific image normalization parameters (mean, standard deviation) beforehand. These parameters are then used by the respective models to standardize all images for both training and inference.

**Supplementary Table 1.** Summary statistics of the processed chest x-ray datasets. The percentage of positive Cardiomegaly cases are kept consistent across training, validation and calibration sets.

|  | All |  | Train |  | Validation |  | Calibration |  | Healthy Adults |  |
| --- | --- | --- | --- | --- | --- | --- | --- | --- | --- | --- |
|  | N | % Positive | N | % Positive | N | % Positive | N | % Positive | N | % Positive |
| **VinDr-PCXR** | 5816 | 0 |  |  |  |  |  |  |  |  |
| **VinDr-CXR** | 14,778 | 14.39% | 9795 | 14.65% | 3265 | 14.64% | 1452 | 14.67% | 266 | 0 |
| **PadChest** | 29,171 | 18% | 18,077 | 19.61% | 6026 | 19.62% | 2678 | 19.60% | 2390 | 0 |
| **NIH** | 20,754 | 7.20% | 12,710 | 7.94% | 4237 | 7.93% | 1884 | 7.96% | 1923 | 0 |
| **CheXBERT** | 6417 | 36.37% | 4064 | 38.93% | 1355 | 38.89% | 593 | 37.94% | 405 | 0 |

### Perturbation Analysis

To minimize harmful exposure when acquiring chest radiographs, children typically receive lower radiation dosages compared to adults, resulting in images with possibly lower contrast or quality^82,83^. Neural networks may latch onto this low-level difference in pediatric images. In addition to our evaluation, we explored how accounting for these differences between datasets can impact the bias on pediatric patients. Histogram matching is a technique that allows us to adjust the pixel distribution of one image to match the pixel distribution of one or more images. We utilized histogram matching to adjust the pixel distributions of imaged healthy children (aged 0 to 1 years old) in the VinDr-PCXR dataset to match the pixel distributions of images in each of the adult-centric datasets, separately. We varied the strength of the histogram matching across different blending ratios. Additionally, we plotted pixel histograms for each dataset using random samples of 1000 images (**Supplementary Figure 1**).

**Supplementary Figure 1.**
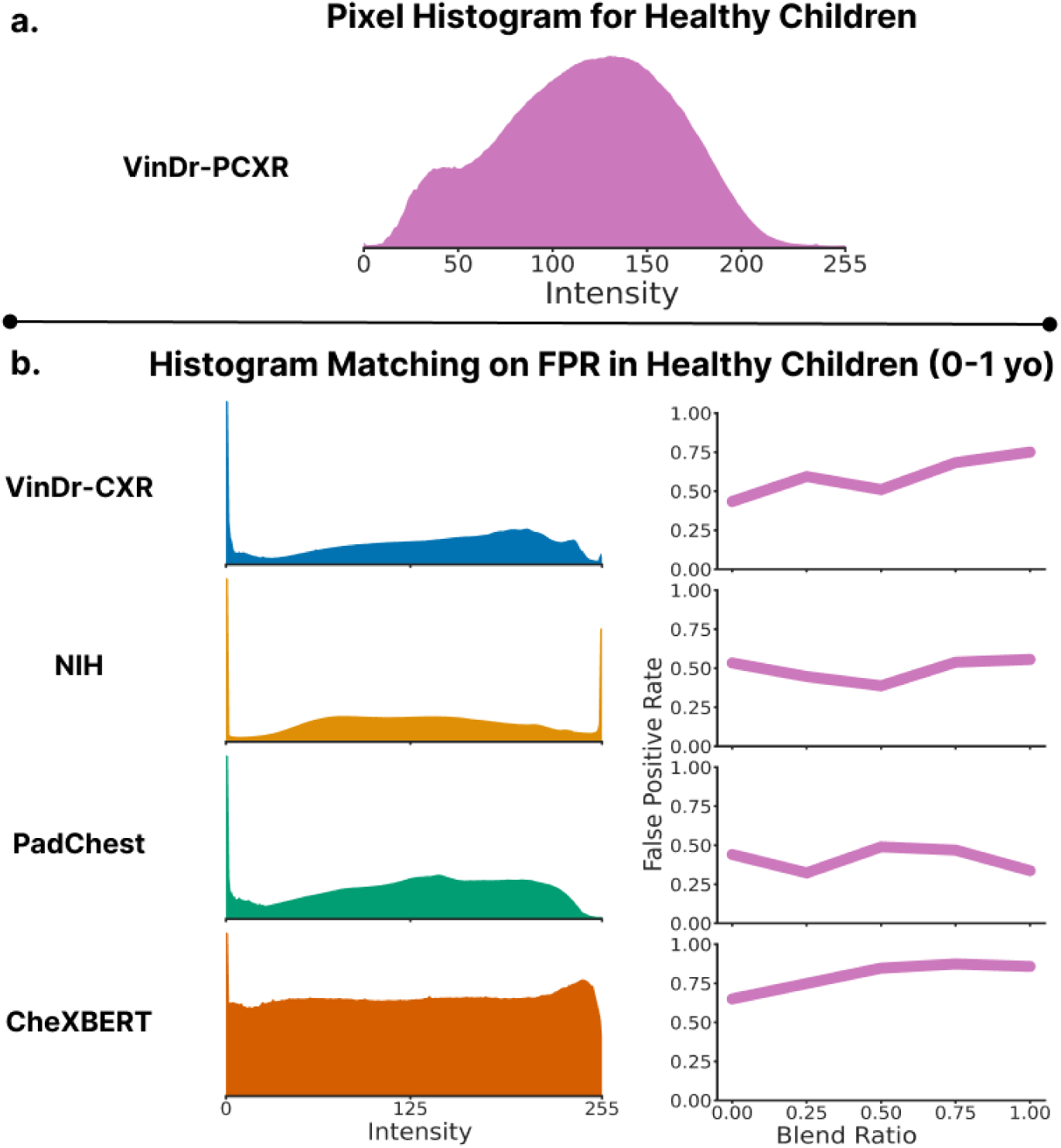
Histogram matching fails to rescue bias against healthy children. **(a)** VinDr-PCXR pixel histogram. **(b)** Pixel histograms for adult datasets **(left)** and impact of histogram matching on the model trained on the corresponding adult dataset **(right).**

## Notes

### Competing Interest Statement

Alex X. Lu is an employee of and holds equity in Microsoft. Alexander J. Towbin receives author royalties from Elsevier, consults for Applied Radiology and previously received funding from Bayer. Irene Y. Chen consults for the Massachusetts Attorney General Office and Flatiron Health.

### Funding Statement

This study did not receive any funding.

### Author Declarations

This paper reviews publicly available datasets that are readily available online. We cite the sources of the public datasets in the paper. Upon publication, we will release a metadata file that specifies all datasets used in the study.

### Summary of Updates

Author institutions have been fixed.

